# Subclinical Vascular Composites Predict Clinical Cardiovascular Disease, Stroke, and Dementia: The Multi-Ethnic Study of Atherosclerosis (MESA)

**DOI:** 10.1101/2023.05.01.23289364

**Authors:** Timothy M. Hughes, Jordan Tanley, Haiying Chen, Christopher L. Schaich, Joseph Yeboah, Mark A. Espeland, Joao A. C. Lima, Bharath Ambale-Venkatesh, Erin D. Michos, Jingzhong Ding, Kathleen Hayden, Ramon Casanova, Suzanne Craft, Stephen R. Rapp, José A. Luchsinger, Annette L. Fitzpatrick, Susan R. Heckbert, Wendy S. Post, Gregory L. Burke

**Author notes:** <u>Corresponding author:</u> Timothy Hughes, PhD, Wake Forest University School of Medicine, Medical Center Blvd, Winston-Salem, NC 27157.

## Abstract

**Background:** Subclinical cardiovascular disease (CVD) measures may reflect biological pathways that contribute to increased risk for coronary heart disease (CHD) events, stroke, and dementia beyond conventional risk scores.

**Methods:** The Multi-Ethnic Study of Atherosclerosis (MESA) followed 6,814 participants (45-84 years of age) from baseline in 2000-2002 to 2018 over 6 clinical examinations and annual follow-up interviews. MESA baseline subclinical CVD procedures included: seated and supine blood pressure, coronary calcium scan, radial artery tonometry, and carotid ultrasound. Baseline subclinical CVD measures were transformed into z-scores before factor analysis to derive composite factor scores. Time to clinical event for all CVD, CHD, stroke and ICD code-based dementia events were modeled using Cox proportional hazards models reported as area under the curve (AUC) with 95% Confidence Intervals (95%CI) at 10 and 15 years of follow-up. All models included all factor scores together and adjustment for conventional risk scores for global CVD, stroke, and dementia.

**Results:** After factor selection, 24 subclinical measures aggregated into four distinct factors representing: blood pressure, arteriosclerosis, atherosclerosis, and cardiac factors. Each factor significantly predicted time to CVD events and dementia at 10 and 15 years independent of each other and conventional risk scores. Subclinical vascular composites of arteriosclerosis and atherosclerosis best predicted time to clinical events of CVD, CHD, stroke, and dementia. These results were consistent across sex and racial and ethnic groups.

**Conclusions:** Subclinical vascular composites of arteriosclerosis and atherosclerosis may be useful biomarkers to inform the vascular pathways contributing to events of CVD, CHD, stroke, and dementia.

## Introduction

Cardiovascular risk summarizes important modifiable and non-modifiable risk factors for cardiovascular disease (CVD), which includes coronary heart disease (CHD) and stroke, that extend to the risk of dementia, and even dementia-related neuropathology.^1-6^ Clinical CVD risk scores applied to dementia are generally summarized by combinations of conventional clinical CVD risk factors into risk scores, and may include education, physical inactivity, and apolipoprotein epsilon 4 (*APOE*-ε4) status.^7,8^ These can include shared or divergent risk factors summarized together as clinical risk factor scores, polygenic risk scores,^1^ and social determinants of health,^2^ which may relate to the biomarkers and risk for dementia.^3,6^ CVD risk scores are intended to scale risk for complex clinical endpoints, such as all CVD, CHD, and stroke; yet, they don’t provide direct measurement of the underlying physiology which may link vascular disorders to CVD and dementia in late-life. Further, most conventional vascular risk scores were originally derived from cohorts of mostly European descent; therefore, these may not be generalizable to other racial and ethnic groups and may misrepresent risk assessment in the general population. As a result, knowledge regarding the impact of vascular disorders on cognitive health in diverse populations is non-specific and limited. We propose herein that composites of subclinical CVD aggregate into dissociable physiologic constructs that represent distinct forms of underlying CVD pathophysiology and may clarify the pathways through which vascular disorders relate to cardiac events, stroke, and dementia. Our goal is not to develop new risk scores with clinical utility. Instead, we seek to develop physiologic composites of vascular disorders. Compared to conventional clinical risk factors, subclinical vascular composites may be more informative biomarkers of the specific pathways underlying the vascular events and the vascular contributions to cognitive impairment and dementia, including Alzheimer’s disease and age-related disorders.

The longitudinal Multi-Ethnic Study of Atherosclerosis (MESA) provides a uniquely large repertoire of subclinical cardiovascular assessments in a racially, ethnically, and regionally diverse group of adults initially free from clinical CVD at baseline. We leveraged these data to create novel composite factor scores collected at baseline by factor analysis and related these subclinical composite factor scores to incident events of all CVD, CHD, stroke, and dementia, adjusted for conventional clinical risk scores for CVD, stroke, and dementia.

## Methods

### Study population

MESA is comprised of 6,814 adults aged 45-84 years free from clinical CVD at baseline who self-reported their race and ethnicity as White, Black, Hispanic, or Chinese at the baseline examination in 2000-2002.^4^ MESA participants were recruited from six areas in the US: Baltimore, Maryland; Chicago, Illinois; Forsyth County, North Carolina; Los Angeles County, California; Northern Manhattan and the Bronx, New York; and Saint Paul, Minnesota. Informed consent was obtained from each participant at baseline and updated at each examination. Approval was received at each site from the local institutional review board for each examination.

### Demographic, Clinical, and Subclinical Measurements

At the baseline examination (Exam 1, 2000-2002), standardized questionnaires were used to collect data on participants’ medical history and demographics including age, education, race and ethnicity. Standard vascular risk scores for each MESA participant were calculated using published equations including the Atherosclerotic Cardiovascular Disease – Pooled Cohort Equation (ASCVD-PCE),^9^ Framingham Global CVD risk score (FRS),^10^ Framingham 10-year Stroke Risk Score (FRS-Stroke),^11^ and Cardiovascular Risk Factors, Aging, and Incidence of Dementia (CAIDE)^12^ drawn from questionnaire and clinical examination during the MESA Exam 1 visit as previously described.^5^ Subclinical cardiovascular assessments at baseline included: carotid ultrasound for carotid plaque, adventitial diameter, and intima-media thickness;^13,14^ coronary artery calcium score by cardiac computed tomography;^15^ augmentation index, large and small artery elasticity, total vascular impedance, estimated cardiac output, stroke volume, and radial tonometry.^16^ Cardiac MRI and brachial flow mediated dilation were only completed on a subset of participants at the baseline examination. *APOE* isoforms were estimated from single nucleotide polymorphisms rs429358 and rs7412 from the genotyping conducted in all MESA participants.

### Ascertainment of clinical events

Telephone interviews were conducted every 9-12 months to inquire about interim hospital admissions and deaths. Copies of death certificates and corresponding International Classification of Diseases (ICD) 10th version codes, face sheets and ICD codes from hospital records and cardiovascular outpatient diagnoses were assembled at each center. For all CVD events, including CHD and stroke events, the full medical records were requested and reviewed and adjudicated by the MESA Morbidity and Mortality committee.^13^ All CHD events were defined as non-fatal events of angina, myocardial infarction, resuscitated cardiac arrest, as well as fatal cardiovascular events of CHD and other fatal CVD. Stroke was classified as present or absent and consisted of rapid onset of a documented focal neurologic deficit lasting 24 hours or until death, or if < 24 hours, there was a clinically relevant lesion on brain imaging. Patients with focal neurologic deficits secondary to brain trauma, tumor, infection, or other non-vascular cause were excluded. Strokes were subclassified on the basis of neuroimaging or other tests into subarachnoid hemorrhage, intraparenchymal hemorrhage, other hemorrhage, brain infarction, or other stroke.^17^ Methodology for identification of incident dementia by ICD codes at hospitalization or death and its validity was previously reported in MESA.^18^ Breifly, a set of ICD-codes used to identify dementia. Dementia as characterized by a significant decline in cognitive function compared to a previous level, not accounted for by other mental disorders (such as major depressive disorder, schizophrenia) or secondary conditions (due to either infection, malignancy, trauma, or substance use). One clinician read medical records blinded to ICD codes, looking for phrases that would indicate, or contradict the conditions defined above.

### Statistical Analysis

Of the 6,814 participants at MESA Exam 1, 98 individuals did not complete at least 3 subclinical cardiovascular procedures necessary to derive factor scores, an additional 29 individuals were missing time to event data for CVD, CHD, stroke, and dementia and were removed from the analytic sample leaving 6,687 for the analysis. Factor analysis initially included 31 subclinical cardiovascular physiologic measures available at Exam 1. Of these, 24 measures (**Supplementary Table 1**) remained after factor selection criteria. Factor selection eliminated subclinical cardiovascular measures that: 1) were obtained on less than 75% of participants at baseline (e.g., cardiac MRI and brachial flow mediated dilation), 2) were highly collinear with other measures (Pearson correlation coefficient >0.90) within another factor (e.g., seated blood pressure measured on different devices), and 3) contributed factor loading <0.25 to each factor. Prior to clustering, each subclinical measure was z-transformed. Principal axis factoring method was used for factor extraction.^19^ The final number of factors was chosen based on eigenvalues and the Scree plot. Factor loadings after Varimax rotation were used to assist with the interpretation of underlying factors. Subclinical composite factors were created using a least square regression approach. A second confirmatory analytic approach use hierarchical agglomerative clustering to sort the subclinical cardiovascular measures into groups based on a bottom up manner and Ward’s minimum variance method was used to arrange the clusters by minimizing the within-cluster variance, producing more compact clusters.^20^

Cox proportional hazards models were used to determine the hazard ratios and 95% confidence intervals (HR, 95% CI) for each subclinical composite factor score adjusted for all other composite factor scores and standardized z-scores for conventional risk factor scores (ASCVD-PCE, FRS, FRS-Stroke, and CAIDE) for each event type (all CVD, CHD, stroke, and ICD-based dementia). The model assumptions were checked using Schoenfeld residuals and Kaplan Meier Curves. Time-dependent Receiver Operating Curve (ROC) Analysis was conducted to determine the individual contributions of subclinical composite factors and conventional clinical risk scores for each outcome. The area under the ROC curve (AUC) at years 10 and 15 were computed for each model, comparing each individual composite factor’s AUC relative to the conventional clinical risk factor score AUC using the method of inverse probability of censoring weighting conditioned on demographics **(Table 1)** to account for censoring in the calculation of AUC. Difference in AUC between each model was reported along with 95% CI. Finally, we assessed potential effect modification for CVD prediction by age, gender, race/ethnicity, and *APOE*-ε4 carrier status.

**Table 1:** Baseline characteristics for MESA participants by event type.

| Characteristics | Statistics | Participants Included |  | Coronary Heart Disease Events |  | Stroke Events |  | ICD-Defined Dementia Events |  |
| --- | --- | --- | --- | --- | --- | --- | --- | --- | --- |
|  |  | (n=6687) |  | (n=684) |  | (n=299) |  | (n=527) |  |
| Age, years | mean, std | 62 | 10 | 66 | 10 | 68 | 10 | 73 | 8 |
| Women | n,% | 3525 | 53 | 246 | 36 | 145 | 48 | 260 | 49 |
| Race/Ethnicity | n,% |  |  |  |  |  |  |  |  |
| White | n,% | 2588 | 39 | 283 | 41 | 111 | 37 | 243 | 46 |
| Chinese | n,% | 800 | 12 | 72 | 11 | 24 | 8 | 34 | 6 |
| Black | n,% | 1853 | 28 | 179 | 26 | 84 | 28 | 147 | 28 |
| Hispanic | n,% | 1475 | 22 | 150 | 22 | 80 | 27 | 103 | 20 |
| Height, cm | mean, std | 166 | 10 | 168 | 10 | 166 | 10 | 165 | 10 |
| Weight, lb | mean, std | 173 | 38 | 178 | 38 | 173 | 37 | 168 | 33 |
| BMI categories | n,% |  |  |  |  |  |  |  |  |
| Normal | n,% | 1937 | 29 | 164 | 24 | 74 | 25 | 150 | 28 |
| Overweight-I | n,% | 2635 | 39 | 289 | 42 | 127 | 42 | 222 | 42 |
| Overweight-II | n,% | 1906 | 28 | 204 | 30 | 88 | 29 | 144 | 27 |
| Overweight-III | n,% | 238 | 4 | 27 | 4 | 10 | 3 | 11 | 2 |
| Education >HS | n, % | 4261 | 64 | 413 | 60 | 170 | 57 | 286 | 54 |
| Systolic BP (mmHg) | mean, std | 127 | 21 | 134 | 22 | 137 | 23 | 136 | 23 |
| Diastolic BP (mmHg) | mean, std | 72 | 10 | 74 | 11 | 74 | 11 | 72 | 10 |
| Cholesterol | mean, std | 194 | 36 | 194 | 39 | 195 | 34 | 193 | 33 |
| Diabetes | n,% |  |  |  |  |  |  |  |  |
| Normal | n,% | 4932 | 74 | 426 | 62 | 186 | 62 | 348 | 66 |
| Impaired fasting glucose | n,% | 916 | 14 | 101 | 15 | 44 | 15 | 90 | 17 |
| Untreated diabetes | n,% | 179 | 3 | 29 | 4 | 9 | 3 | 16 | 3 |
| Treated diabetes | n,% | 668 | 10 | 127 | 19 | 58 | 19 | 72 | 14 |
| Cholesterol medication use | n,% | 1081 | 16 | 159 | 23 | 57 | 19 | 111 | 21 |
| Antihypertensive use | n,% | 2223 | 33 | 313 | 46 | 149 | 50 | 249 | 47 |
| APOE ε4** | n,% |  |  |  |  |  |  |  |  |
| Missing | n,% | 423 | 6 | 53 | 8 | 15 | 5 | 25 | 5 |
| No ε4 alleles | n,% | 4590 | 69 | 471 | 69 | 222 | 74 | 331 | 63 |
| 1 or 2 ε4 allele(s) | n,% | 1674 | 25 | 160 | 23 | 62 | 21 | 171 | 32 |
| ASCVD-PCE (% risk) | mean, std | 14% | 13% | 21% | 15% | 22% | 16% | 26% | 15% |
| FRS (% risk) | mean, std | 14% | 10% | 21% | 9% | 20% | 9% | 21% | 9% |
| FSRS (% risk) | mean, std | 5% | 5% | 7% | 6% | 8% | 7% | 10% | 7% |
| CAIDE | mean, std | 8 | 3 | 9 | 3 | 9 | 2 | 9 | 2 |

Blood pressure (BP), Atherosclerotic Cardiovascular Disease – Pooled Cohort Equation (ASCVD-PCE), Framingham Global CVD risk score, Framingham 10-year Stroke Risk Score (FSRS), Cardiovascular Risk Factors, Aging, and Incidence of Dementia (CAIDE)

## Results

At Exam 1, 6,687 MESA participants had a mean (SD) age of 62 (10) years ranging between 45-85 years, 53% were women, 13% had diabetes, and 33% were taking blood pressure medications (**Table 1**). Over a maximum of 17 years of follow-up, the median [interquartile range] of follow-up time to first event was 15.6 [10.4, 16.5] years. A total of 1,023 participants had all CVD events, including 684 CHD events and 299 stroke events, and 527 had a death or hospitalization with an ICD code for dementia. Compared to the entire cohort, individuals with clinical events were more likely to be older, men, have a lower education, and higher conventional risk scores at baseline.

Subclinical cardiovascular measures clustered into four composite factors (**Supplemental Table 1**), representing: blood pressure (**Factor_BP** from systolic, diastolic, mean arterial pressure, systolic vessel diameter on ultrasound), atherosclerosis (**Factor_Athero** from maximum carotid stenosis, coronary artery calcification, common and internal carotid intima media thickness), arteriosclerosis (**Factor_Arterio** by ankle brachial index, pulse pressure, distensibility coefficient, Young’s Elastic Modulus, pulse wave reflection magnitude and aortic augmentation index, large and small artery elasticity, systemic vascular resistance, and total vascular impedance), and cardiac function (**Factor_Cardiac** by pulse rate, pulse pressure amplification, estimated cardiac output, estimated ejection time, and estimated stroke volume). These subclinical composite factors representing blood pressure, atherosclerosis, arteriosclerosis, and cardiac function showed minimal to modest relationships with each other (**Supplemental Figure 1)**. Results from the factor analysis were compared with the results from hierarchical clustering using the wardD.2 method to confirm composite grouping (**Supplemental Figure 2**). Subclinical composite factors showed moderate correlations with conventional clinical cardiovascular risk scores (**Supplemental Table 2**). Factor_BP was most consistently correlated with all conventional clinical risk scores: ASCVD (rho = 0.32), Framingham Risk Score (*r* = 0.37), FRS-Stroke (*r* = 0.45), and CAIDE risk score (*r* = 0.32). Factor_Athero, representing atherosclerosis was most correlated with ASCVD (*r* = 0.47) and Framingham Risk Score (*r* = 0.46). While Factor_Arterio was most strongly correlated with ASCVD-PCE (*r* = 0.30) and Factor_Cardiac was most strongly correlated with FRS-Stroke (*r* = 0.33), these composite factor scores were minimally correlated with other conventional risk scores. Subclinical composite factors for atherosclerosis and arteriosclerosis were positively associated with age (**Supplemental Figure 3**).

**Table 2** presents the proportional hazards of subclinical composite factors and time-to-event for CHD, stroke, and ICD-defined dementia events when all subclinical composite factors were considered together in the same model and a second model that also included one of the conventional clinical risk scores. We observed that each subclinical composite factor was independently associated with decreased time to each event in models for CHD, stroke, and dementia events. After adjustment for conventional risk scores, each subclinical composite factor remained significantly associated with time to events, showing independent contributions to event prediction beyond conventional clinic risk scores and the presence of the other subclinical composite factors. We observed similar associations with events when each composite factor was considered alone (data not shown).

**Table 2.**
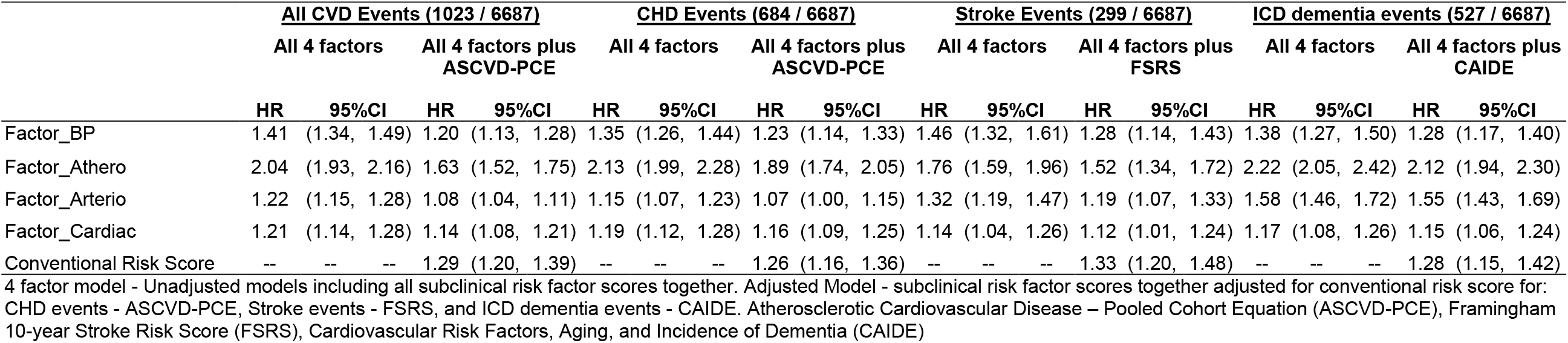
Proportional hazards models of subclinical vascular composites and incident events (n=6687).

Time to event prediction models for CHD, stroke, and ICD-defined dementia events where constructed for each subclinical vascular composite factor alone relative to conventional risk scores and all subclinical vascular composite factors with conventional risk scores at 10 and 15 years of follow-up. The 10-year event range is consistent with the methods used to develop conventional CVD and stroke prediction risk scores and is highlighted in **Figure 1. Supplemental Figure 4** shows the predictive ability for each subclinical vascular composite factors and conventional risk scores corresponding to 15 years of follow-up. Each subclinical composite factor alone provided significant yet modest prediction for all events independent of each other and conventional clinical risk scores. The addition of subclinical composite factors significantly improved AUC by contributing to the prediction models of CVD events, CHD events, and ICD-defined dementia events beyond established risk scores. Of note, Factor_Athero alone predicted CVD events at 10 years of follow-up nearly as well as conventional risk scores composites of known clinical risk factors, as evidenced by non-significant differences in AUC differences, as indicated in **Table 3**. These differences were no longer significant at 15 years of follow-up. Further, Factor_Athero and Factor_Arterio alone provided similar or better prediction of time to ICD-defined dementia events at 10 years of follow-up as CAIDE which includes key known risk factors for dementia (e.g., age, education, *APOE*-ε4). This similarity in predictive ability of CAIDE and Factor_Arterio continued to 15 years of follow-up. Factor_Athero provided significantly better prediction of ICD-defined dementia events than CAIDE at 10 and 15 years of follow-up. The addition of subclinical factor scores improved model prediction of CVD and ICD-defined dementia events over 10 and 15 years of follow-up, but did not improve prediction of stroke events.

**Figure 1.**
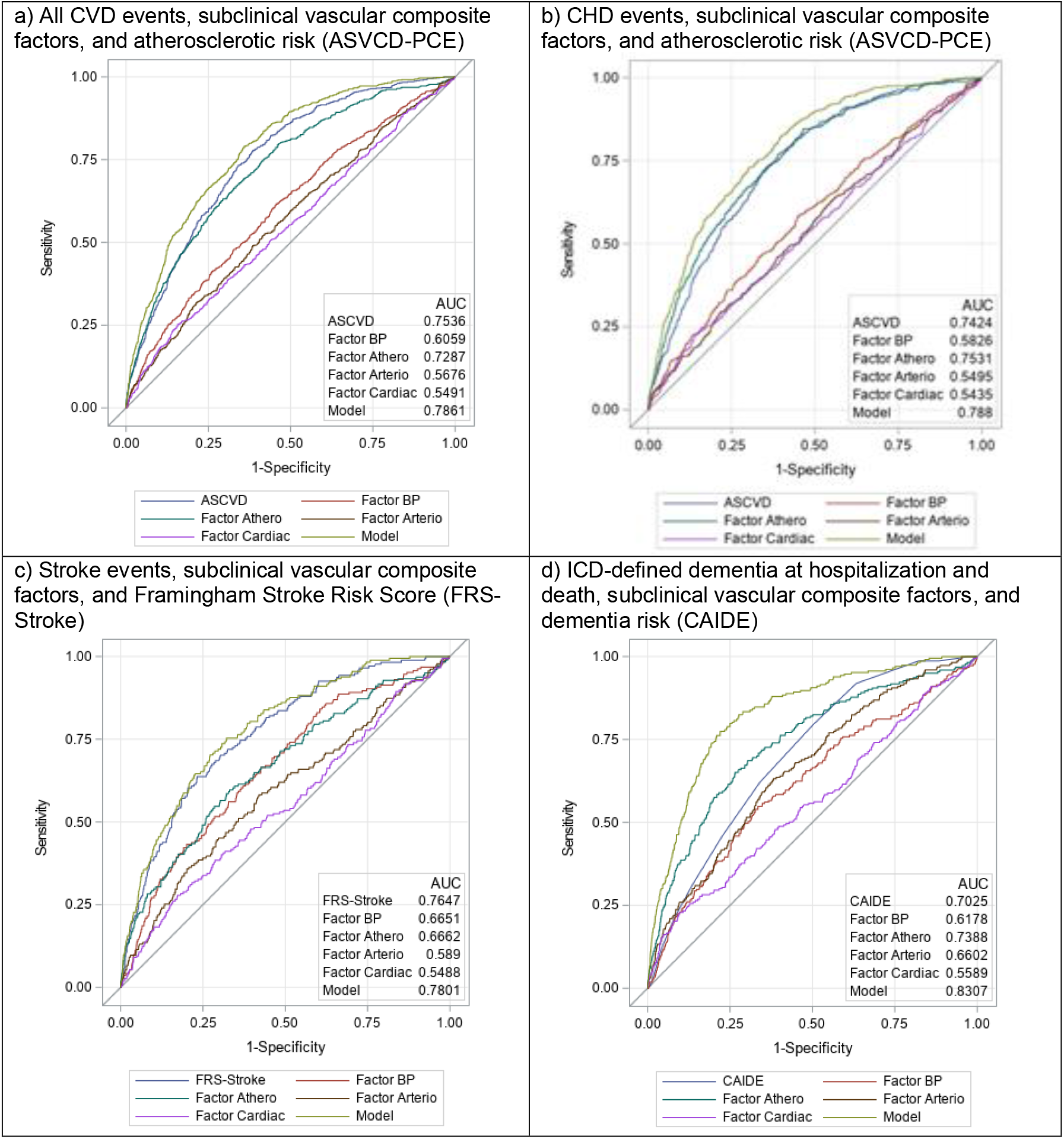
ROC curves for subclinical composite factors and events of all cardiovascular disease (CVD), coronary heart disease (CHD), stroke, and ICD-defined dementia at 10 years. Cardiovascular Risk Factors, Aging, and Incidence of Dementia (CAIDE). Model refers to all predictors together, including all composite factors and the conventional risk score.

**Table 3.** AUC differences for each subclinical composite factor relative to AUC for conventional clinical risk score alone in ROC analysis.

| Effect | 10 Year Estimates |  |  | 15 Year Estimates |  |  |
| --- | --- | --- | --- | --- | --- | --- |
|  | AUC | (95% CI) |  | AUC | (95% CI) |  |
|  | Difference^ |  |  | Difference^ |  |  |
| <u>All Cardiovascular Events (1023 / 6687)</u> |  |  |  |  |  |  |
| Factor_BP | -0.15 | (-0.12, -0.17) | * | -0.17 | (-0.16, -0.19) | * |
| Factor_Athero | -0.02 | (0.00, -0.05) | * | -0.06 | (-0.04, -0.08) | * |
| Factor_Arterio | -0.19 | (-0.16, -0.21) | * | -0.19 | (-0.17, -0.22) | * |
| Factor_Cardiac | -0.20 | (-0.17, -0.24) | * | -0.22 | (-0.19, -0.25) | * |
| ASCVD 10 year | Referent (AUC = 0.754) |  |  | Referent (AUC = 0.777) |  |  |
| <u>Coronary Heart Disease Events (684 / 6687)</u> |  |  |  |  |  |  |
| Factor_BP | -0.16 | (-0.13, -0.19) | * | -0.17 | (-0.15, -0.19) | * |
| Factor_Athero | 0.01 | (0.04, -0.01) |  | -0.02 | (0.01, -0.04) |  |
| Factor_Arterio | -0.19 | (-0.16, -0.23) | * | -0.20 | (-0.18, -0.23) | * |
| Factor_Cardiac | -0.20 | (-0.16, -0.24) | * | -0.21 | (-0.18, -0.24) | * |
| ASCVD 10 year | Referent (AUC =0.742) |  |  | Referent (AUC = 0.763) |  |  |
| <u>Stroke Events (299 / 6687)</u> |  |  |  |  |  |  |
| Factor_BP | -0.10 | (-0.05, -0.15) | * | -0.15 | (-0.11, -0.19) | * |
| Factor_Athero | -0.10 | (-0.06, -0.13) | * | -0.10 | (-0.08, -0.13) | * |
| Factor_Arterio | -0.18 | (-0.13, -0.22) | * | -0.16 | (-0.12, -0.20) | * |
| Factor_Cardiac | -0.22 | (-0.16, -0.27) | * | -0.23 | (-0.19, -0.27) | * |
| FSRS 10 year | Referent (AUC = 0.765) |  |  | Referent (AUC = 0.773) |  |  |
| <u>ICD-Defined Dementia Events (527 / 6687)</u> |  |  |  |  |  |  |
| Factor_BP | -0.08 | (-0.04, -0.13) | * | -0.10 | (-0.07, -0.13) | * |
| Factor_Athero | 0.04 | (0.08, 0.00) |  | 0.06 | (0.09, 0.03) | * |
| Factor_Arterio | -0.04 | (0.01, -0.09) |  | -0.03 | (0.01, -0.06) |  |
| Factor_Cardiac | -0.14 | (-0.09, -0.20) | * | -0.12 | (-0.09, -0.15) | * |
| CAIDE | Referent (AUC = 0.703) |  |  | Referent (AUC = 0.690) |  |  |

All models include subclinical composite factor scores adjusted for conventional clinical risk factor score.
^indicates estimates and p-values relative to conventional clinical risk score. \*indicates significant difference (at p-value <0.05) relative to the conventional risk score. Non-significant differences denote where subclinical factors show equivalent AUC as conventional risk scores. Significant differences denote where subclinical factors show less predictive ability than the conventional risk scores. Conventional risk scores include: Atherosclerotic Cardiovascular Disease – Pooled Cohort Equation (ASCVD-PCE, 10 year), Framingham 10-year Stroke Risk Score (FSRS, 10-year), Cardiovascular Risk Factors, Aging, and Incidence of Dementia (CAIDE)

After evaluating potential interactions between each subclinical composite factor score and age, race and ethnicity, gender, and *APOE*-ε4 in the all CVD events model, no significant interactions were detected between factor scores and gender, race and ethnicity, or with *APOE*-ε4 (p-interaction all >0.05). Significant interactions were observed between factor scores and baseline age with respect to CVD events, such that factor scores significantly predicted CVD events in both participants <60 and ≥60 years at baseline; yet, factor scores tended to perform best among participants <60 years of age at baseline.

## Discussion

Creation of subclinical vascular composite factors using a large repository of subclinical cardiovascular measures collected at baseline in a large, multi-ethnic, and longitudinal cohort showed that these measures aggregate into four discrete primary factors representing blood pressure, atherosclerosis, arteriosclerosis, and cardiac function. The resulting subclinical vascular composite factors were dissociable. They showed low correlations with each other and modest correlations with conventional clinical vascular risk scores. Each subclinical composite factor independently predicted time to CVD events, CHD, and stroke events, as well as ICD-defined dementia events over 17 years of follow-up. These results were largely consistent after adjustments for conventional clinical CVD risk scores that summarize established risk factors for CVD and dementia. Prediction models for both CHD and ICD-defined dementia events clearly showed that the addition of subclinical vascular factors improved risk prediction models. While subclinical vascular composite factors were significantly associated with stroke events, their addition to the conventional clinic risk (e.g., FRS-Stroke) did not significantly improve stroke prediction. The observed relationships with all CVD events appear to be stronger among participants <60 years of age at baseline. We did not observe significantly effect modification by gender or race/ethnicity in this diverse cohort suggesting that these relationships between subclinical vascular risk factors and events appear to be consistent across subgroups at differential risk for CVD events and dementia. Further, the benefit of quantifying subclinical vascular factors (atherosclerosis, arteriosclerosis, blood pressure, cardiac function) significantly improved the prediction of all CVD and dementia events independent of conventional clinical risk scores in this cohort. These findings provide important and clinically meaningful results that suggests targeted control of subclinical vascular disease (e.g. blood pressure, arterial stiffness, atherosclerosis) may further reduce the risk for clinical events for CVD and dementia.

These results suggest that subclinical vascular composites representing aspects of vascular pathology are significant predictors of incident events of CVD and ICD-defined dementia over 17 years of follow-up. These relationships are consistent across gender, *APOE*-ε4 genotype and racial/ethnic groups, but prediction of CVD events was significantly stronger among adults <60 years of age. The factor representing atherosclerosis was most significantly associated with all CVD events, including CHD and stroke, as well as dementia events. This is consistent with prior work in MESA showing coronary artery calcification score was an important predictor of CHD and all atherosclerotic cardiovascular disease combined outcomes.^21,22^ The novel factor representing arteriosclerosis was the second most predictive subclinical factor for ICD-defined dementia events. This finding is consistent with prior studies showing that markers of arterial stiffness are strongly associated with cerebral small vessel disease and Alzheimer’s disease pathology.^23,24^ The factor representing blood pressure showed consistently modest relationships with each outcome, supporting evidence that elevated blood pressure plays a basic and consistent role leading to clinical events of CHD, stroke, and dementia. These findings extend recent work from our group which showed that these same conventional clinical risk scores for CHD, stroke, and CAIDE were significantly associated with cross sectional cognitive performance, cognitive decline over 6 years, as well as, lower cortical thickness, greater white matter hyperintensity burden, and amyloid deposition on neuroimaging in this MESA cohort.^5,6^ Taken together, this work suggests the potential research utility of including subclinical pathways of vascular risk in future work looking at the vascular contributions to cognitive impairment and dementia.

Previous studies have evaluated the relationships between subclinical cardiovascular disease composites and survival,^25^ and CVD events^26^ using a limited number of subclinical measures to define an index of: ankle-arm index, electrocardiogram, and common carotid intima-media thickness, based on clinical cutoffs as well as abdominal aortic aneurysms and infarction (>3mm) on brain MRI in a subsample of participants. They found modest hazard ratios for cardiovascular events and mortality within 8 years. In contrast to prior work, our approach intentionally did not combine conventional risk factors with subclinical vascular measures nor did it focus on developing new risk scores with clinical utility. Instead, we developed novel subclinical composite factors representing specfic pathways to underlying vascular disorders and show they improve risk prediction for events.

We recognize that few studies have the combination of detailed subclinical vascular phenotyping and longitudinal assessments necessary to evaluate multiple subclinical vascular risk scores and their relationship to clinical events of CHD, stroke, and dementia over nearly two decades of follow-up. Even fewer studies are multi-ethnic and collect these measures in the middle-age to late-life transition period when vascular factors are expected to begin affecting cerebrovascular health. The MESA cohort was designed to provide extensive subclinical vascular assessments at baseline and longitudinal follow-up of CHD, stroke, and dementia necessary to examine and validate these relationships in a diverse and representative cohort.

When combined, these subclinical composite factor scores in MESA create a unique resource for examining the relative contribution of vascular disorders to late-life CVD and cerebrovascular health. This work is solely intended to development novel constructs representing biologic pathways in the subclinical vascular contributions to heart disease and brain health. These will play an important role in the ongoing work in MESA to examine the vascular contributions to Alzheimer’s disease and related dementias.

We acknowledge a potential point of conceptual contention is the use of the terms designating these biomarkers as “subclinical” measures of atherosclerosis or arteriosclerosis, rather than measures of “arterial injury” or “arteriopathy”.^27^ The “subclinical” factors derived from this work are not intended to facilitate clinical diagnosis of CVD, contribute to develop prediction models, guide clinical practice, nor are they intended to be directly transferable to other less phenotyped cohorts or patients. These initial composite factors provide a level of resolution on vascular disorders we have not seen in prior studies and may shed light on mechanistic features of each subclinical cardiovascular phenotype. As we have shown here, these factors provide a physiologic construct representing dissociable pathways of biologic cardiovascular disorders that increase the risk for CHD, stroke, and dementia events. The current approach only considers baseline measures available in nearly all participants. Additional work will be needed to create reference ranges and weighting for individual factors represented in MESA should this new information be adapted for repeated measures or clinical use. The use of ICD codes at hospitalization and death for dementia in this analysis are not optimal approaches to examine the Alzheimer’s disease and related dementias. Passive surveillance and the reliance on clinical diagnostic codes for dementia is neither sensitive nor specific for cognitive impairment and dementia subtyping. Future studies will be needed to examine the relationships between subclinical vascular factors, cognitive adjudication of cognitive impairment, and imaging and plasma biomarkers of Alzheimer’s disease and related dementias. We propose that subclinical vascular composite factors may be more useful biomarkers than conventional dementia risk scores to interrogate the specific vascular contributions to cognitive impairment and dementia including Alzheimer’s disease related dementias, and have the potential to illuminate the vascular pathways potentially underlying these age-related disorders.

Vascular composite factors created from multiple aspects of subclinical cardiovascular measures representing atherosclerosis, arteriosclerosis, blood pressure, and cardiac function demonstrate that higher subclinical vascular burden is associated with time to CHD events, stroke events, and a clinical diagnosis of dementia over 17 years of follow-up. Subclinical factors of arteriosclerosis and atherosclerosis provide similar or better risk estimation as conventional risk factors for CHD and dementia events and also point to more specific subclinical vascular pathways to CVD events and dementia that deserve to be investigated further in mechanistic studies.

## Data Availability

Data are available upon request from the Multi-Ethnic Study of Atherosclerosis.

http://www.mesa-nhlbi.org

---

This research was supported by contracts 75N92020D00001, HHSN268201500003I, N01-HC-95159, 75N92020D00005, N01-HC-95160, 75N92020D00002, N01-HC-95161, 75N92020D00003, N01-HC-95162, 75N92020D00006, N01-HC-95163, 75N92020D00004, N01-HC-95164, 75N92020D00007, N01-HC-95165, N01-HC-95166, N01-HC-95167, N01-HC-95168 and N01-HC-95169 from the National Heart, Lung, and Blood Institute, by grants UL1-TR-000040, UL1-TR-001079, and UL1-TR-001420 from the National Center for Advancing Translational Sciences (NCATS), and grants R01AG054069 and R01AG058969 from the National Institute on Aging. The authors thank the other investigators, the staff, and the participants of the MESA study for their valuable contributions. A full list of participating MESA investigators and institutions can be found at http://www.mesa-nhlbi.org. This paper has been reviewed and approved by the MESA Publications and Presentations Committee.

**Supplemental Table 1.**
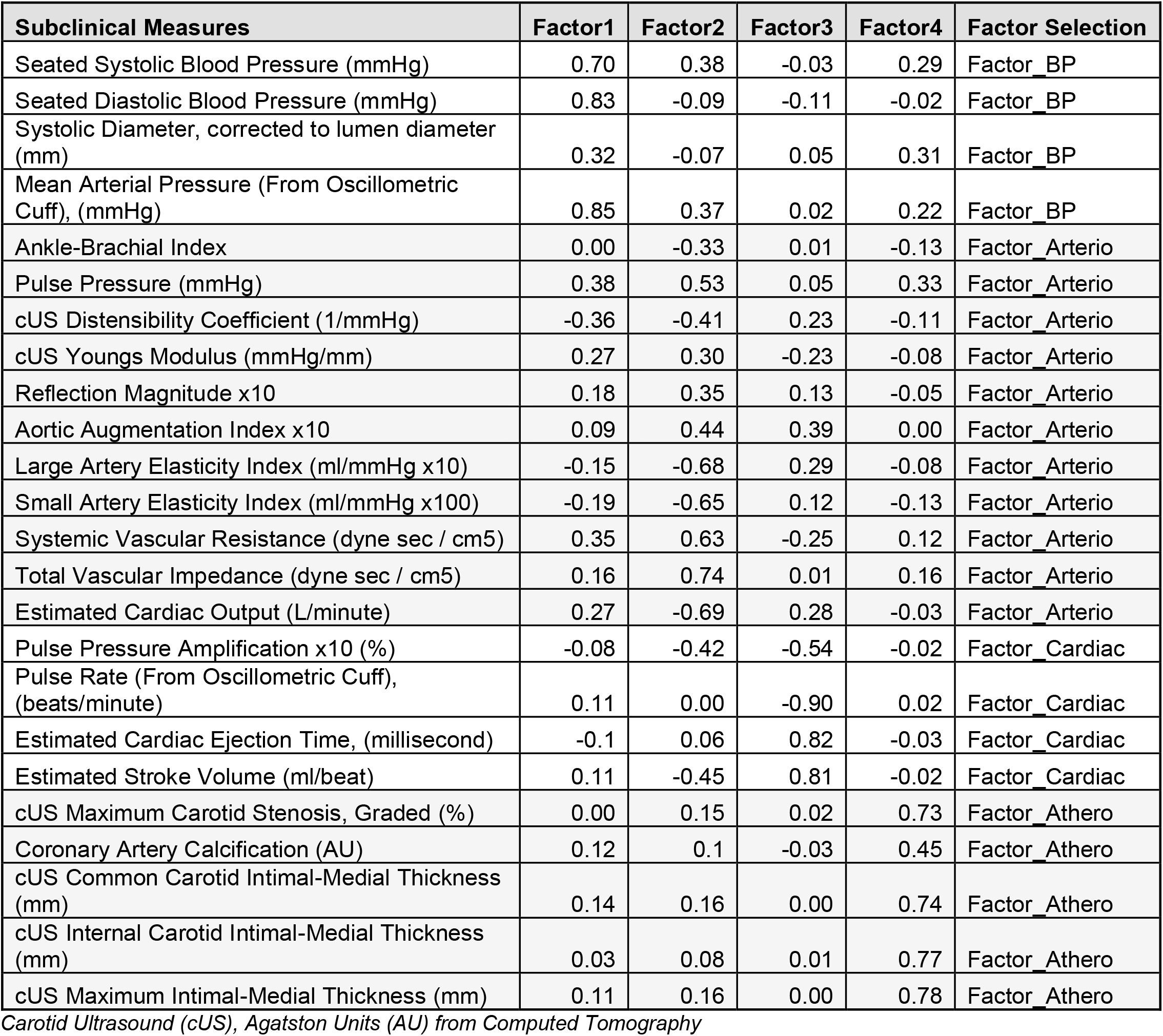
Subclinical Cardiovascular Measures, Factor Loading Scores, and Factor Selection from MESA Baseline Examination.

| Subclinical Measures | Factor1 | Factor2 | Factor3 | Factor4 | Factor Selection |
| --- | --- | --- | --- | --- | --- |
| Seated Systolic Blood Pressure (mmHg) | 0.70 | 0.38 | -0.03 | 0.29 | Factor_BP |
| Seated Diastolic Blood Pressure (mmHg) | 0.83 | -0.09 | -0.11 | -0.02 | Factor_BP |
| Systolic Diameter, corrected to lumen diameter (mm) | 0.32 | -0.07 | 0.05 | 0.31 | Factor_BP |
| Mean Arterial Pressure (From Oscillometric Cuff), (mmHg) | 0.85 | 0.37 | 0.02 | 0.22 | Factor_BP |
| Ankle-Brachial Index | 0.00 | -0.33 | 0.01 | -0.13 | Factor_Arterio |
| Pulse Pressure (mmHg) | 0.38 | 0.53 | 0.05 | 0.33 | Factor_Arterio |
| cUS Distensibility Coefficient (1/mmHg) | -0.36 | -0.41 | 0.23 | -0.11 | Factor_Arterio |
| cUS Youngs Modulus (mmHg/mm) | 0.27 | 0.30 | -0.23 | -0.08 | Factor_Arterio |
| Reflection Magnitude x10 | 0.18 | 0.35 | 0.13 | -0.05 | Factor_Arterio |
| Aortic Augmentation Index x10 | 0.09 | 0.44 | 0.39 | 0.00 | Factor_Arterio |
| Large Artery Elasticity Index (ml/mmHg x10) | -0.15 | -0.68 | 0.29 | -0.08 | Factor_Arterio |
| Small Artery Elasticity Index (ml/mmHg x100) | -0.19 | -0.65 | 0.12 | -0.13 | Factor_Arterio |
| Systemic Vascular Resistance (dyne sec / cm5) | 0.35 | 0.63 | -0.25 | 0.12 | Factor_Arterio |
| Total Vascular Impedance (dyne sec / cm5) | 0.16 | 0.74 | 0.01 | 0.16 | Factor_Arterio |
| Estimated Cardiac Output (L/minute) | 0.27 | -0.69 | 0.28 | -0.03 | Factor_Arterio |
| Pulse Pressure Amplification x10 (%) | -0.08 | -0.42 | -0.54 | -0.02 | Factor_Cardiac |
| Pulse Rate (From Oscillometric Cuff), (beats/minute) | 0.11 | 0.00 | -0.90 | 0.02 | Factor_Cardiac |
| Estimated Cardiac Ejection Time, (millisecond) | -0.1 | 0.06 | 0.82 | -0.03 | Factor_Cardiac |
| Estimated Stroke Volume (ml/beat) | 0.11 | -0.45 | 0.81 | -0.02 | Factor_Cardiac |
| cUS Maximum Carotid Stenosis, Graded (%) | 0.00 | 0.15 | 0.02 | 0.73 | Factor_Athero |
| Coronary Artery Calcification (AU) | 0.12 | 0.1 | -0.03 | 0.45 | Factor_Athero |
| cUS Common Carotid Intimal-Medial Thickness (mm) | 0.14 | 0.16 | 0.00 | 0.74 | Factor_Athero |
| cUS Internal Carotid Intimal-Medial Thickness (mm) | 0.03 | 0.08 | 0.01 | 0.77 | Factor_Athero |
| cUS Maximum Intimal-Medial Thickness (mm) | 0.11 | 0.16 | 0.00 | 0.78 | Factor_Athero |

*Carotid Ultrasound (cUS), Agatston Units (AU) from Computed Tomography*

**Supplemental Table 2.** Pearson correlation coefficients for subclinical vascular composite factor and clinical risk scores.

|  | **ASCVD-**<br>**PCE** | **FRS** | **FRS-**<br>**Stroke** | **CAIDE** |
| --- | --- | --- | --- | --- |
| **Factor_BP** | 0.316 | 0.371 | 0.450 | 0.320 |
| **Factor_Athero** | 0.474 | 0.461 | 0.305 | 0.270 |
| **Factor_Arterio** | 0.299 | 0.159 | 0.108 | 0.194 |
| **Factor_Cardiac** | 0.149 | 0.170 | 0.333 | 0.126 |

All $p < 0.05$ . Atherosclerotic Cardiovascular Disease – Pooled Cohort Equation (ASCVD-PCE), Framingham Global CVD risk score (FRS), Framingham 10-year Stroke Risk Score (FSRS), Cardiovascular Risk Factors, Aging, and Incidence of Dementia (CAIDE).

**Supplemental Figure 1:**
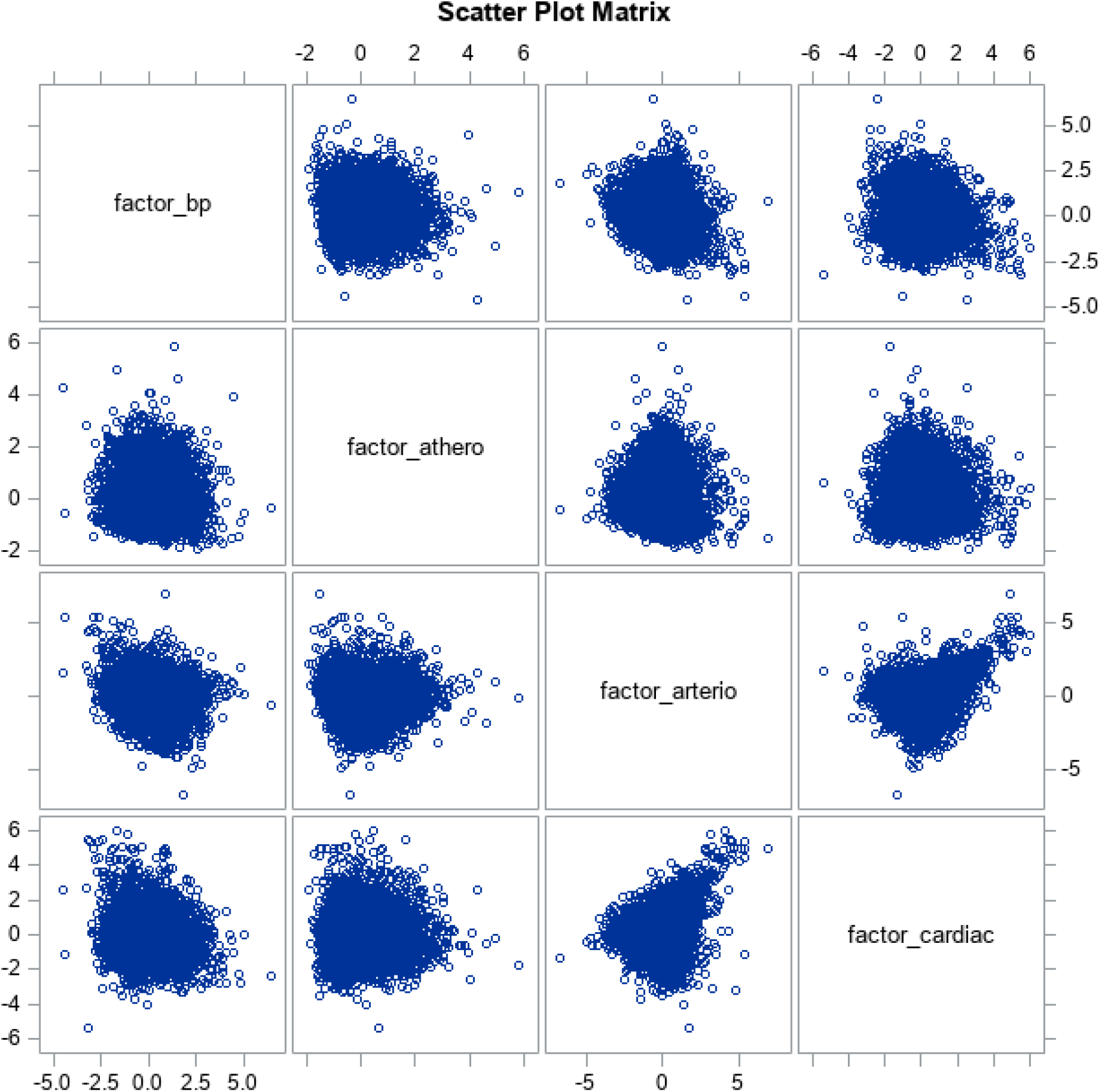
Scatterplot matrix of derived subclinical composite factors of blood pressure, atherosclerosis, arteriosclerosis, and cardiac function.

**Supplemental Figure 2.**
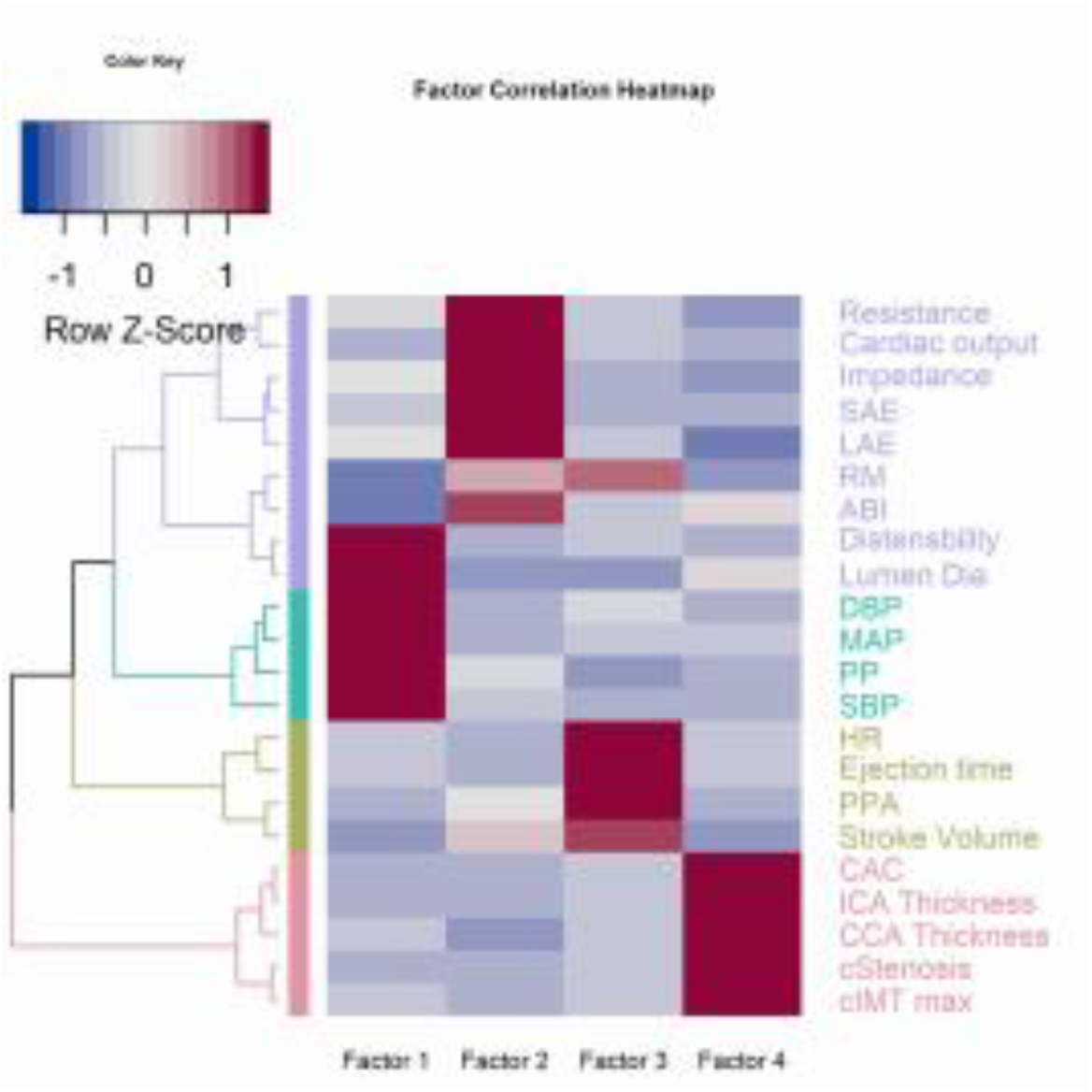
Heatmap and row dendrogram of ward.D hierarchical clustering method. Hierarchical clustering of repertoire of subclinical vascular biomarkers measured at MESA Exam 1 (2000-02). The heatmap visualizes a data matrix of the subclinical vascular biomarkers (rows) and factors (columns). The tree-like diagram on the left of the heatmap is a dendrogram which orders the biomarkers by similarity. Hierarchical agglomerative clustering sorted the subclinical vascular biomarkers into groups based on a bottom-up manner. The different colors highlight the four different clusters formed. Ward’s minimum variance method (ward.D) was used to arrange the clusters by minimizing the within-cluster variance, producing more compact clusters.

**Supplemental Figure 3.**
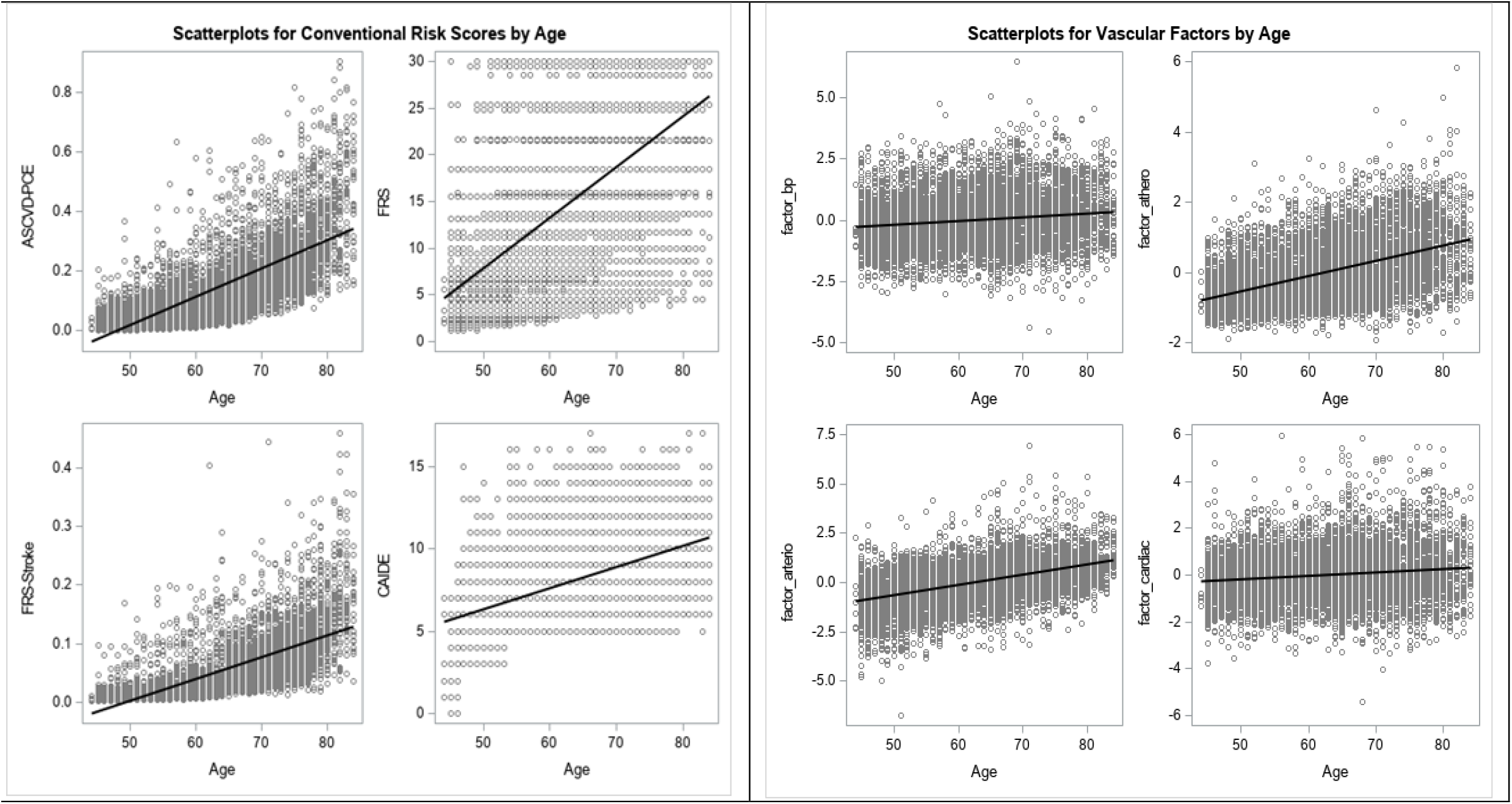
Scatterplots of subclinical cardiovascular composite factor scores and conventional clinical risk scores by age at MESA baseline. Lines represent linear regession with age. Atherosclerotic Cardiovascular Disease – Pooled Cohort Equation (ASCVD-PCE), Framingham Global CVD risk score (FRS), Framingham 10-year Stroke Risk Score (FSRS), Cardiovascular Risk Factors, Aging, and Incidence of Dementia (CAIDE).

**Supplemental Figure 4.**
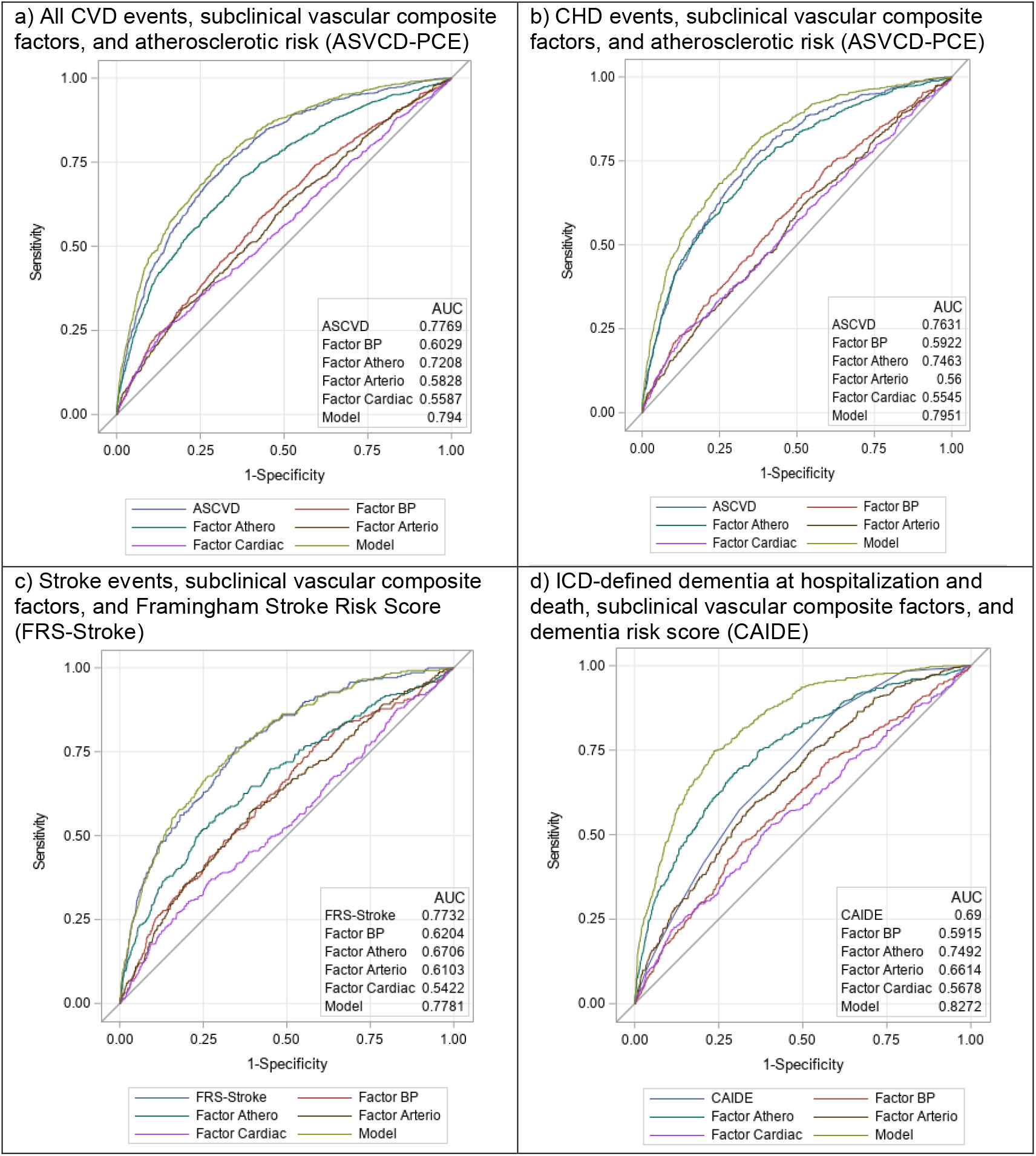
ROC curves for subclinical vascular composite factor scores and time to events of all cardiovascular disease, coronary heart disease, stroke, and ICD-defined dementia at 15 years. Cardiovascular Risk Factors, Aging, and Incidence of Dementia (CAIDE).

